# Development and Pilot Implementation of a Classification for Hemodialysis Cannulation Infiltrations: A Single-Center Quality Improvement Study

**DOI:** 10.64898/2026.11.26.26368860

**Authors:** Vandana Dua Niyyar, Forest Rawls, Rodella Broxton-Key, Derrick F. Jones

## Abstract

**Background:** Cannulation-related complications, including infiltrations, are inconsistently defined and documented, making it difficult to establish reliable baselines and compare cannulation techniques and technologies, and assess improvements in care. Further, existing classifications do not systematically capture downstream clinical consequences.

**Methods:** To address these gaps, we developed a novel grading classification for infiltrations, combining immediate local injury severity with modifiers for downstream clinical consequences. The classification was piloted in a quality improvement project at Emory outpatient hemodialysis units from July 1 through September 30, 2026. All infiltrations during the study period were recorded and assigned both severity grades and clinical modifiers; management and follow-up were summarized descriptively.

**Results:** Among 12 reported infiltrations, seven were recorded as Grade 1, three as Grade 2, and two as Grade 3. Dialysis was disrupted in 11 treatments, with access rest required in 9 patients. In 6 instances, a pre-existing central venous catheter was used until the arteriovenous access was able to be successfully reused. Among 11 episodes with documented reuse, the median time to reuse was 7 days (range, 0–44 days).

**Conclusions:** The classification supports systematic documentation of infiltration severity and clinically relevant consequences, providing a starting point for establishing baselines and tracking cannulation outcomes. Further refinement and validation are needed before it can support comparisons across settings or evaluation of improvements in dialysis access care.

## Introduction

Vascular access complications are significant contributors to hospitalizations and health care costs among patients receiving hemodialysis [1,2]. A fundamental, and often overlooked, determinant of vascular access durability is cannulation success. Despite the unignorable fact that sustained, successful use of an arteriovenous fistula or graft depends on reliable cannulation, with two needle insertions required at each hemodialysis treatment, cannulation-related complications remain understudied [3,4]. In addition to immediate sequelae and impact on patient experience, infiltrations may have significant long-term downstream consequences that affect access survival. A landmark study demonstrated that major fistula infiltrations were associated with new or prolonged catheter dependence, additional diagnostic studies, surgical evaluations or procedures, and fistula thrombosis [5]. Though this study was conducted >20 years ago, it remains relevant even today as we have yet to establish a baseline for frequency of infiltrations and their consequences.

To establish reference standards that could serve as a starting point for measuring outcome improvements, standardized definitions and systematic reporting of cannulation-related complications are needed [6] and can be built upon currently existing frameworks. The Kidney Health Initiative’s work on vascular access clinical trial endpoints [7] can be expanded to incorporate cannulation-specific outcomes. The KDOQI classification of infiltration injuries as minor, major, or severe [8] establishes the groundwork for a more detailed and specific construct. Building on these, we developed a novel grading classification to document infiltrations, incorporating the extent of the local injury and DIRECT modifiers for clinically relevant consequences including dialysis treatment disruption, troubleshooting, access rest, prolongation of catheter dependence, need for emergent referral and subsequent interventions (Tables 1a and 1b). This report describes its initial three-month clinical application and priorities for further evaluation.

**Table 1a.** Grading Classification for Cannulation-Related Infiltrations.

| **Grade** | **Severity** | **Clinical findings** |
|---|---|---|
| **1** | Mild | Local swelling, bruising, or hematoma measuring <2 cm in maximum dimension. |
| **2** | Moderate | Local swelling, bruising, or hematoma measuring 2–10 cm, inclusive. |
| **3** | Severe | Local swelling, bruising, or hematoma measuring >10 cm. |
| **4** | Catastrophic | Rapidly expanding hematoma, compartment syndrome, or new distal neurovascular compromise attributable to the infiltration. |

At initial assessment, measure the longest dimension across the visible or palpable injury and record the measurement time. Update the grade if reassessment identifies a higher-grade finding. Grade 4 takes precedence regardless of size.

**Table 1b.** DIRECT Modifiers for Cannulation-Related Infiltration. Record all applicable modifiers independently of the injury grade.

| **Code** | **Description** |
|---|---|
| **D** | Dialysis paused, shortened, or missed due to the infiltration. Record whether the prescribed treatment was completed. |
| **I** | Additional access evaluation or intervention. Record referrals, diagnostic testing completed, and therapeutic procedures performed separately. |
| **R** | Access rest required due to the infiltration. Record its duration. |
| **E** | Emergency department evaluation or emergency hospital admission due to the infiltration. |
| **C** | Central venous catheter use required due to the infiltration. Specify whether an existing catheter was used or a new one was placed. |
| **T** | Troubleshooting: Needle repositioning or recannulation due to the infiltration. |

AVF, arteriovenous fistula. AVG, arteriovenous graft. CVC, central venous catheter.

## Methods

We piloted a novel infiltration classification in a quality improvement initiative at Emory outpatient hemodialysis units from July 1 through September 30, 2026. Clinical teams and nursing managers reported cannulation-related infiltrations to the Vascular Access Team (FR, RB, DJ), who collected event details, assigned severity grades, and tracked subsequent clinical consequences. When the team was off-site at the time of infiltration, they relied on the information provided by the dialysis staff on the floor. The proposed framework grades infiltration severity from mild to catastrophic and uses modifiers for disrupted dialysis treatment through the access (D), troubleshooting including needle repositioning (T), requirement for access rest (R), need for central venous catheter use (C), emergent referral (E) and subsequent interventions (I) (Tables 1a and 1b).

For the descriptive analysis, age was reported in non-overlapping 10-year age groups. Time from access creation to first use was calculated using the date of first two-needle cannulation recorded in the patient’s chart. Time to access reuse was calculated from the infiltration date to the first recorded date of subsequent successful two-needle cannulation.

## Results

Twelve infiltrations were reported during the three-month pilot (Table 2). Median age was 54.5 years (range, 41–89 years), and there were 5 males and 7 females. Nine infiltrations were recorded in arteriovenous fistulas (AVF) and three in arteriovenous grafts (AVG). 17-gauge needles were used in two patients, 16-gauge in nine episodes, and 15-gauge in one. The median interval from access creation to first recorded two-needle cannulation was 73 days (range, 35–228 days).

**Table 2.** Characteristics of the 12 Cannulation Infiltration Episodes.

| Episode | Age Range<sup>a</sup> (years) | Sex | Access | Needle gauge | Grade | Severity | DIRECT codes<sup>b</sup> | Creation to first use<sup>c</sup> (days) | Infiltration to reuse<sup>d</sup> (days) |
|---|---|---|---|---|---|---|---|---|---|
| 1 | 70-79 | Male | BC AVF | 16 | 1 | Mild | T | 73 | 0 |
| 2 | 50-59 | Female | BBT AVF | 17 | 2 | Moderate | D, I, R, C | 172 | 44 |
| 3 | 40-49 | Male | RC AVF | 17 | 1 | Mild | D, R, C | 59 | 42 |
| 4 | 50-59 | Male | RC AVF | 16 | 1 | Mild | D, I, R, C | 38 | 17 |
| 5 | 40-49 | Female | BC AVF | 16 | 1 | Mild | D, R | 145 | 2 |
| 6 | 40-49 | Female | RC AVF | 16 | 1 | Mild | D, R, C | 228 | Pt refused |
| 7 | 50-59 | Male | RC AVF | 16 | 3 | Severe | D, I, R, C | 73 | 42 |
| 8 | 80-89 | Male | AVG | 16 | 1 | Mild | D, T | 35 | 3 |
| 9 | 80-89 | Female | AVG | 16 | 2 | Moderate | D, T | 64 | 2 |
| 10 | 40-49 | Female | RC AVF | 16 | 2 | Moderate | D, R | 130 | 3 |
| 11 | 60-69 | Female | AVG | 16 | 1 | Mild | D, R, C | 56 | 8 |
| 12 | 40-49 | Female | BC AVF | 15 | 3 | Severe | D, I, R, T | 84 | 7 |

Abbreviations. AVF, arteriovenous fistula. AVG, arteriovenous graft. BBT, transposed brachiobasilic. BC, brachiocephalic. CVC, central venous catheter. RC, radiocephalic.
<sup>a</sup> Age at pilot initiation is reported in non-overlapping 10-year bands.
<sup>b</sup> Grades and modifiers are from the spreadsheet’s final classification. D, dialysis disruption. I, additional access evaluation or intervention. R, access rest. E, emergency evaluation or admission. C, catheter use. T, needle troubleshooting. All C codes in this cohort refer to pre-existing CVCs.
<sup>c</sup> First use is the date of first two-needle cannulation recorded in the patient’s chart.
<sup>d</sup> Reuse is the date of first subsequent successful two-needle cannulation use of the infiltrated access. A value of 0 indicates same-day reuse.

Seven episodes were classified as Grade 1 (mild; visible bruising from infiltration < 2 cm from puncture site), three as Grade 2 (moderate; visible bruising from infiltration 2 - 10 cm from puncture site), and two as Grade 3 (severe; visible bruising from infiltration > 10 cm from puncture site). No episode was recorded as Grade 4 (catastrophic).

Every episode had at least one recorded DIRECT modifier (Table 2). The dialysis treatment was disrupted in 11 episodes and troubleshooting with needle repositioning was attempted in 4 patients. The dialysis access was rested in 9 instances and for 6 of these, the patient received dialysis through a pre-existing central venous catheter. Four of the patients required additional access evaluation or intervention including formal ultrasonography, vascular surgery evaluation, and angioplasty. There were no infiltrations requiring emergency management.

Clinical consequences varied within grades. One Grade 1 episode was managed by recannulation at another site after a brief delay, with successful access reuse on the same day. Four other Grade 1 episodes involved use of a pre-existing catheter. In one of these, the access was reused after self-cannulation training, and in another, after a surgical review that was already previously scheduled for evaluation of the fistula. One Grade 2 episode involved a patient who did not have a pre-existing central venous catheter and missed dialysis treatments until successful fistula reuse. Both Grade 3 episodes required access rest and further access evaluation or intervention. One involved use of an existing catheter with subsequent angioplasty of multiple stenotic lesions. The other involved early termination of dialysis, needle repositioning, and subsequent angioplasty of a cephalic arch stenosis.

Among the 11 episodes with documented reuse, the median interval from infiltration to first subsequent successful two-needle cannulation was 7 days (range, 0–44 days). In the remaining Grade 1 episode, the fistula remained unused at last documentation because of patient preference who had been initially reluctant to use her access, and the infiltration further strengthened her reluctance.

## Discussion

We developed this classification to capture two critical aspects of cannulation-related infiltration: the extent of injury observed in the dialysis unit and its subsequent impact on access outcomes. To make bedside grading objective and reproducible, we based it on the maximum dimension of swelling, bruising, or hematoma as Grade 1 (<2 cm, localized to the puncture site), Grade 2 (2–10 cm, inclusive), or Grade 3 (>10 cm). Catastrophic events, including rapidly expanding hematoma, compartment syndrome, or new neurovascular compromise, were designated Grade 4 regardless of size. Separate modifiers captured subsequent clinically relevant consequences as described above. These distinctions are essential as a relatively small (but deep) infiltration can disrupt dialysis or require catheter use, while extensive bruising, particularly in frail older patients with thinning skin, may look striking yet have little effect on treatment or subsequent access use. Additionally, the term infiltration, as it is used currently, covers a wide range of events, from needle misplacement that can be addressed by repositioning and trouble-shooting at one end of the spectrum, to a catastrophic injury followed by access loss at the other end of the spectrum. A designation such as for example Grade 2 (D, R, T) gives dialysis teams a shared description of the episode: a mild local injury accompanied by dialysis disruption, access rest, and needle troubleshooting.

This single-center pilot was limited by its small sample size and short observation period. Local staffing and access availability may also have influenced how infiltrations were managed. When a pre-existing catheter is available, using it may be easier than troubleshooting cannulation, particularly when the vascular access team is unavailable to assist. Catheter use after infiltration may therefore reflect practical considerations as well as injury severity. As the appearance and extent of bruising may evolve after the initial assessment, the grade assigned at the time of infiltration may underestimate the maximum extent of injury. Additionally, when the vascular access team was off-site, episodes were reported after the event. We intend to integrate the classification into our electronic medical record and expand its use by all staff across our dialysis units to support longitudinal tracking including photographs taken on the day of infiltration to document the initial appearance. Longer follow-up and multicenter validation are needed to assess reproducibility and applicability across settings.

The classification captures staff-assessed injury and documented clinical consequences; future iterations should incorporate the patient experience as well. Qualitative studies describe anticipatory anxiety and disrupted sleep related to needle-related distress, as well as the importance of feeling safe and trusting the cannulator [9,10]. A Kidney Health Initiative workgroup found that the impact of vascular access on quality of life is poorly characterized in trials, registries, and quality programs and recommended standardized use of access-specific patient-reported outcome measures [11]. Revisions and improvement of similar classifications could include direct patient input and integrate injury grading with a validated experience measure such as the 28-item Needling Patient Reported Experience Measure (NPREM) [12].

Further, adoption of these or similar classifications by LDOs (large dialysis organizations) encouraging standardized, nonpunitive reporting at the point of care could complement claims data by linking each documented infiltration to its immediate and subsequent consequences. With cannulation denominators, dialysis programs could establish baseline rates and characterize injury severity, treatment disruption, and other clinical consequences. Claims analyses cannot determine whether cannulation trauma contributed to later stenosis or thrombosis, so these events should be tracked separately and interpreted in the right clinical context [6]. These measures could help assess whether changes in cannulation practice or new technologies, such as routine bedside ultrasound, reduce the frequency or clinical impact of infiltrations [13]. Patterns of events, interpreted in the context of access complexity and cannulation volume, could also help identify staff who would benefit from additional training and the skills to target. Future studies could evaluate whether simulation-based practice, direct observation, and structured feedback improve cannulation outcomes [14]. Pairing these measures with patient-reported experience would show whether improvements in technical performance are accompanied by improvements patients notice and value.

## Conclusion

This classification provides a starting point for developing a common lexicon to document hemodialysis cannulation infiltrations and their clinical consequences. We invite dialysis teams to pilot the framework and share their experience to help refine its definitions, grades, and modifiers across practice settings. Our hope is that adoption and consistent use of such a framework in the future would help dialysis programs establish reliable baselines, compare cannulation practices, and determine whether quality improvement interventions lead to better outcomes.

## Data Availability

All data produced in the present study are available upon reasonable request to the authors

## Acknowledgments

The authors would like to thank Prince Niyyar for his suggestion to create a memorable acronym for the clinical modifiers.

## References

1. United States Renal Data System. 2025 USRDS Annual Data Report: Epidemiology of Kidney Disease in the United States. End-stage renal disease. Chapter 4: Vascular access. National Institute of Diabetes and Digestive and Kidney Diseases. https://usrds-adr.niddk.nih.gov/2025/end-stage-renal-disease/4-vascular-access

2. Thamer M, Lee TC, Wasse H, et al. Medicare costs associated with arteriovenous fistulas among US hemodialysis patients. Am J Kidney Dis. 2018;72(1):10–18. doi:10.1053/j.ajkd.2018.01.034.

3. van Loon MM, Kessels AGH, van der Sande FM, Tordoir JHM. Cannulation and vascular access-related complications in hemodialysis: factors determining successful cannulation. Hemodial Int. 2009;13(4):498–504. doi:10.1111/j.1542-4758.2009.00382.x.

4. Kumbar L, Ramani K, Brouwer-Maier D. Considerations in access cannulation: traditional and evolving approaches. Adv Chronic Kidney Dis. 2020;27(3):199–207. doi:10.1053/j.ackd.2020.03.007.

5. Lee T, Barker J, Allon M. Needle infiltration of arteriovenous fistulae in hemodialysis: risk factors and consequences. Am J Kidney Dis. 2006;47(6):1020–1026. doi:10.1053/j.ajkd.2006.02.181.

6. Gage SM, Reichert H. Determining the incidence of needle-related complications in hemodialysis access: we need a better system. J Vasc Access. 2021;22(4):521–532. doi:10.1177/1129729820946917.

7. Beathard GA, Lok CE, Glickman MH, et al. Definitions and end points for interventional studies for arteriovenous dialysis access. Clin J Am Soc Nephrol. 2018;13(3):501–512. doi:10.2215/CJN.11531116.

8. Lok CE, Huber TS, Lee T, et al. KDOQI clinical practice guideline for vascular access: 2019 update. Am J Kidney Dis. 2020;75(4 Suppl 2):S1–S164. doi:10.1053/j.ajkd.2019.12.001.

9. Duncanson EL, Chur-Hansen A, Le Leu RK, et al. Dialysis needle-related distress: patient perspectives on identification, prevention, and management. Kidney Int Rep. 2023;8(12):2625–2634. doi:10.1016/j.ekir.2023.09.011.

10. Fielding C, Bramley L, Stalker C, Brand S, Toft S, Buchanan H. Patients’ experiences of cannulation of arteriovenous access for haemodialysis: a qualitative systematic review. J Vasc Access. 2023;24(5):1121–1133. doi:10.1177/11297298211067630.

11. Woo K, Fuld R, Grandinetti A, et al. Patient-reported outcomes in hemodialysis vascular access: a call to action. J Vasc Access. 2022;23(6):973–980. doi:10.1177/11297298211018295.

12. Moore C, Busby A, Flanagan R, et al. Development and validation of a measure to assess patient experience of needling of arteriovenous fistulas or grafts for haemodialysis access: the NPREM. Clin Kidney J. 2025;18(3):sfaf029. doi:10.1093/ckj/sfaf029.

13. Niyyar VD, Buch K, Rawls F, Broxton R. Effectiveness of ultrasound-guided cannulation of AVF on infiltration rates: a single center quality improvement study. J Vasc Access. 2023;24(2):322–328. doi:10.1177/11297298211034280.

14. Singapogu R, Chowdhury A, Roy-Chaudhury P, Brouwer-Maier D. Simulator-based hemodialysis cannulation skills training: a new horizon? Clin Kidney J. 2021;14(2):465–470. doi:10.1093/ckj/sfaa206.

